# Trends across time in socioeconomic differences in body mass index: a comparison of population and individual-level approaches

**DOI:** 10.1101/2023.05.24.23290477

**Authors:** Liam Wright, Charis Bridger Staatz, Richard J Silverwood, David Bann

**Affiliations:** Centre for Longitudinal Studies, Social Research Institute, University College London

**Keywords:** variation explained, predictive accuracy, body mass index, obesity, social inequalities, social epidemiology

## Abstract

**Background:** Socioeconomic differences in body mass index (BMI) have widened alongside the obesity epidemic. However, the utility of socioeconomic position (SEP) indicators at the individual level remains uncertain, as does the potential temporal variation in their predictive value. Examining this is important in light of the increasing incorporation of SEP indicators into predictive algorithms and the possibility that SEP has become a more important predictor of BMI over time. We thus investigated SEP differences in BMI over three decades of the obesity epidemic in England and compared population-wide (SEP group differences in mean BMI) and individual-level (out-of-sample prediction of individuals’ BMI) approaches.

**Methods:** We used repeated cross-sectional data from the Health Survey for England, 1991-2019. BMI (kg/m^2^) was measured objectively, and SEP was measured via educational attainment and neighborhood index of deprivation (IMD). We ran random forest models for each survey year and measure of SEP adjusting for age and sex.

**Results:** The mean and variance of BMI increased within each SEP group over the study period. Mean differences in BMI by SEP group also increased across time: differences between lowest and highest education groups were 1.0 kg/m^2^ (0.4, 1.6) in 1991 and 1.5 kg/m^2^ (0.9, 1.8) in 2019. At the individual level, the predictive capacity of SEP was low, though increased in later years: including education in models improved predictive accuracy (mean absolute error) by 0.14% (−0.9, 1.08) in 1991 and 1.06% (0.17, 1.84) in 2019. Similar patterns were obtained when analyzing obesity, specifically.

**Conclusion:** SEP has become increasingly important at the population (group difference) and individual (prediction) levels. However, predictive ability remains low, suggesting limited utility of including SEP in prediction algorithms. Assuming links are causal, abolishing SEP differences in BMI could have a large effect on population health but would neither reverse the obesity epidemic nor explain the vast majority of individual differences in BMI.

## Introduction

Obesity rates have more than tripled among adults in England since 1980 [1]. Average body mass index (BMI) has also increased, but the population distribution of BMI has become more spread and more skewed [2], implying that individuals have not been equally affected by the obesity epidemic. Given the substantial health and economic costs associated with obesity [3], identifying solutions to the obesity epidemic continues to be an area of significant policy and research interest.

A large amount of research has focused on social inequalities in obesity and BMI (see, e.g., [4–7] for reviews). Recent evidence finds that adults in the most deprived areas of England are twice as likely to be obese as those in the least deprived areas [8]; a similar difference is observed comparing highest and lowest education groups [8]. Evidence further suggests that, in England, inequalities in obesity and BMI according to education level have widened – in absolute terms – alongside the development of the obesity epidemic [8–10], a pattern observed in multiple other countries [11], though not all [12].

Research on social inequalities in BMI has typically taken a population level approach and focused on estimating associations – for instance, examining the mean difference in BMI according to educational attainment. Less attention has been paid to the explanatory power of socioeconomic factors at the individual level – for instance, the proportion of between-person variability in BMI that can be predicted by socioeconomic position (SEP) [13]. Though measures of SEP have been included in predictive algorithms for BMI [14] and reducing social inequality has been proposed as a way to tackle high obesity rates [15], SEP appears to explain only a small amount (< 6%) of between-person variability in BMI [12,16–20]. This is the case even when multiple indicators of SEP across life are used [16,17].

The comparatively low explanatory power of SEP accords with more general observations. Twin studies find that the variance in adult BMI explained by environmental factors shared between twins (such as parental SEP) is very low, in contrast to the proportion explained by genetics and non-shared environmental factors [21,22]. This low explanatory power is observed across almost all traits – so much so that it is known as the ‘gloomy prospect’ in behavioural genetics [23–27]. Attempts to directly predict individual life outcomes using SEP and other survey data have produced humbling results. For example, a recent scientific mass collaboration showed that several socioeconomic outcomes were largely unpredictable using a range of sophisticated predictive models and unusually rich survey data (including socioeconomic histories) [28].

While the explanatory power of SEP on BMI may be lower than perhaps expected [15], it could have systematically changed across time. The increasing variation of population BMI partly reflects increasing inequalities *between* SEP groups, but it reflects increasing variation *within* these groups, too [2,18,29–32]. If the increasing variation *within* groups exceeds increasing variation *between* groups, the explanatory power of SEP – already low – may have fallen further still. Determining whether this is the case is important for understanding the role of SEP as a contributor to the obesity epidemic [29] and for understanding the (continuing) potential for using SEP in predictive algorithms. However, research on this question is limited. Studies from the United States [12] and Indonesia [18] find the explanatory power of SEP on BMI has decreased over time, but social inequalities declined in these countries over the periods assessed. Thus, results may not generalize to England or other countries that have experienced widening inequalities across time.

Existing research is further limited by a focus on individual-level (education) and not area-level (e.g., neighbourhood deprivation) measures of SEP – research has highlighted the role of area based factors, such as neighbourhood walkability and fast food outlet density, as contributors to the obesity epidemic [33]. Existing research is also limited by the use of methods not tailored for prediction. In particular, studies have used linear regression models of limited flexibility, which may not have captured interactions and other non-linearities. They have also assessed explanatory power within the same sample as used to estimate models (thus biasing towards more optimistic results) and have not assessed predictive ability (i.e., the difference between predicted and observed BMI), specifically – a metric of particular importance for creating accurate prediction algorithms for BMI.

In this paper, we examined trends in the explanatory and predictive power of individual and area-level SEP on BMI more formally by adopting principles and methods from machine learning. We used random forest models and repeat cross-sectional data from the Health Survey for England (HSE) to examine changes in the predictive ability of educational attainment and neighbourhood deprivation for BMI and obesity between the years 1991-2019, a period in which obesity rates doubled in England [1].

## Methods

### Participants

The HSE is an ongoing series of annual nationally representative cross-sectional health surveys that began in 1991. Detailed description of the survey is available elsewhere [34]. The HSE uses a multi-stage sampling design with households drawn from a list of postcode sectors. Non-response weights are provided with the data from 2002 onwards, due to increasing refusal rates. We used these where available, assuming weights of 1 in other survey years. We limited our analysis to individuals aged 25-64 – the lower bound chosen to focus on ages with few members (1-8%) in full-time education (whose eventual education level is not known) and the upper bound chosen to reduce selection biases that could arise due to higher mortality rates among high BMI individuals [35]. We further limited our sample to those of White ethnicity to create comparable populations less liable to changes in composition due to inflow and outflow of migration. For similar reasons, we also excluded a small number of individuals whose highest qualification was obtained abroad as well as individuals currently in full time education (4.2% of observations). There was only a small amount of missingness on our covariate data (< 0.1%), so we analysed complete cases only. Our final sample size was 145,421. This excluded 10.7% of the eligible sample who had missing BMI data. The sample size each year ranged from 1,813 in 1991 to 9,556 in 1993.

### Measures

#### Body Mass Index

BMI was calculated by dividing weight in kilograms by height in metres squared. Height and weight were measured directly by interviewers. From 1995, individuals weighing more than 130 kg were asked to give an estimate of their weight due to limitations with the scales, so measurements for these individuals are based on self-report.

#### Socioeconomic Position

The HSE contains few measures of SEP that are measured consistently in each wave. We chose to focus on educational attainment and neighbourhood deprivation. Education was recorded using the national vocational qualification schema (NVQ 4/5, higher education below degree level, NVQ 3, NVQ 2, NVQ1, none). Neighbourhood deprivation was measured using the Index of Multiple Deprivation (IMD) and was categorized into quintiles (1^st^ least deprived – 5^th^ most deprived). The IMD combines deprivation across seven domains (income, employment, education, health, crime, barriers to housing and services, and living environment). It is available in the HSE at the electoral ward level from 2001-2002 and lower super output area (LSOA) level thereafter (LSOAs each include 400-1200 households). In the HSE, IMD data are available from 2001 only. New versions of the IMD are released intermittently. The IMD2000 is available from 2001-2002, the IMD2004 from 2003-2007, the IMD2007 from 2008-2010, the IMD2010 from 2011-2014, the IMD2015 from 2015-2018 and the IMD2019 in 2019. We use the data as supplied, with no further attempts made to harmonize across years.

#### Covariates

We included age and sex as covariates in our prediction models. Both may confound or moderate the association between education and IMD and BMI [10,36]. Age was available in single years prior to 2015, but only in five-year categories from 2015 onwards. For consistency we randomly imputed single year ages from 2015 onwards. Mean age increased in our sample between 1991-2019 (average age ∼ 43 in 1991 and ∼ 45 in 2019).

### Statistical Analysis

To maximize predictive ability, we used random forest models; known to provide similar or superior predictions to traditional regression approaches in multiple settings [37,38]. Our analysis consisted of fitting random forest models and assessing their predictive accuracy and explanatory power. Random forests are a decision tree-based method in which data are recursively split according to decision rules invoking individual predictor variables (e.g., male or female, age < 45). Decision rules are chosen such that splits minimise heterogeneity in the target variable (here, BMI). To avoid overfitting, random forests use an ensemble approach where the results of multiple decision trees are averaged, with each tree being fit on a subset of predictor variables and a random sample of observations [39,40]. As predictions are generated via successive binary splits, random forests can account for non-linearities or interactions between independent variables (e.g., between age and education) without requiring their explicit parameterization, an advantage here given previously observed differences in social inequalities in BMI between males and females [41], across cohorts [10], and over the life course [10].

We fit a random forest (500 trees) for BMI for each year of data collection and measure of SEP, using SEP, age, and sex as predictor variables. We then extracted model predictions and used these to calculate three metrics of explanatory power and predictive accuracy: variation explained (R^2^), mean absolute error (the difference between observed and predicted BMI), and probability of superiority. (In this setting, the probability of superiority is the probability that among two randomly chosen participants, the participant with the higher predicted BMI score has the higher observed BMI.) Importantly, to avoid overfitting, we generated model predictions using a portion of our data that was not used to estimate the random forest model (procedure explained further below). R^2^ provides a (relative) measure of how well SEP can predict between-person differences, while mean absolute error and probability of superiority provide summaries of how well SEP can predict individuals.

We compared the three metrics to (a) baseline predictions where mean BMI was used and (b) the results of random forest models including only sex and age as predictor variables. We also examined the progression of the magnitude of the association between educational attainment and BMI by using the results of the random forest models to predict mean BMI assuming everyone in the population had the same SEP. We defined the size of the association between SEP and BMI as the difference in predicted population mean BMI for the most advantaged and disadvantaged SEP categories (NVQ 4/5 vs no qualifications for education and highest vs lowest quintile for IMD). To calculate confidence intervals, we used bootstrapping accounting for the complex survey design (Rao & Wu method [42,43], 500 bootstrap samples). For the predictive accuracy and explanatory power metrics, we used the observations not selected within a given bootstrap to generate predictions to avoid overfitting.

As the random forest models were estimated for each year separately, to more easily ascertain trends in (a) the proportion of prediction error explained by each SEP variable and (b) the size of the association between BMI and each SEP variable, we smoothed the bootstrap estimates by regressing estimates upon year splines using generalized additive models (GAMs) – GAMs allow for flexible, smooth non-linear associations between independent and dependent variables. The change in the age variable to five-year categories from 2015 onwards may have artificially increased the relative incremental predictive power of including SEP in models. Consequently, we also ran the GAM models using data only up to 2014 to assess whether trends were observable prior to the change in the data.

We also performed a series of further analyses. First, as social inequalities in BMI are typically found to be stronger among females than males [5], we repeated the analysis having stratified by sex. Second, as obesity (BMI ≥ 30 kg/m^2^) is of particular research and policy interest, we repeated the analysis using obesity as the outcome measure (see Supplementary Information Results S1 for further detail on methods used). Third, as random forests could potentially overfit the data, we repeated the BMI analysis using simple linear regression. In these models, predictors were included as linear (age) or categorical (sex, education, IMD) terms with no interactions included.

The organization used to conduct the HSE changed in 1994. Some previous studies using HSE have accordingly focused on data from 1994 onwards [44,45]. We present results from 1991-2019, but in the text report results from 1994 where results from 1991-1993 depart considerably from those in later years.

## Results

### Descriptive Statistics

There was an increase in the overall mean and variance of BMI and the prevalence of obesity between 1991-2019 (Figures 1a-1c; see also Supplementary Figure S1). Education levels generally increased across time; the proportion of individuals with the highest education level increased from 11.7% in 1991 to 37.2% in 2019 (Supplementary Figure S2). Increasing education levels led to non-linear changes in the variance of the education measure; variance decreased overall between 1991 to 2019 but peaked in 2002 (Figure 1d).

**Figure 1:**
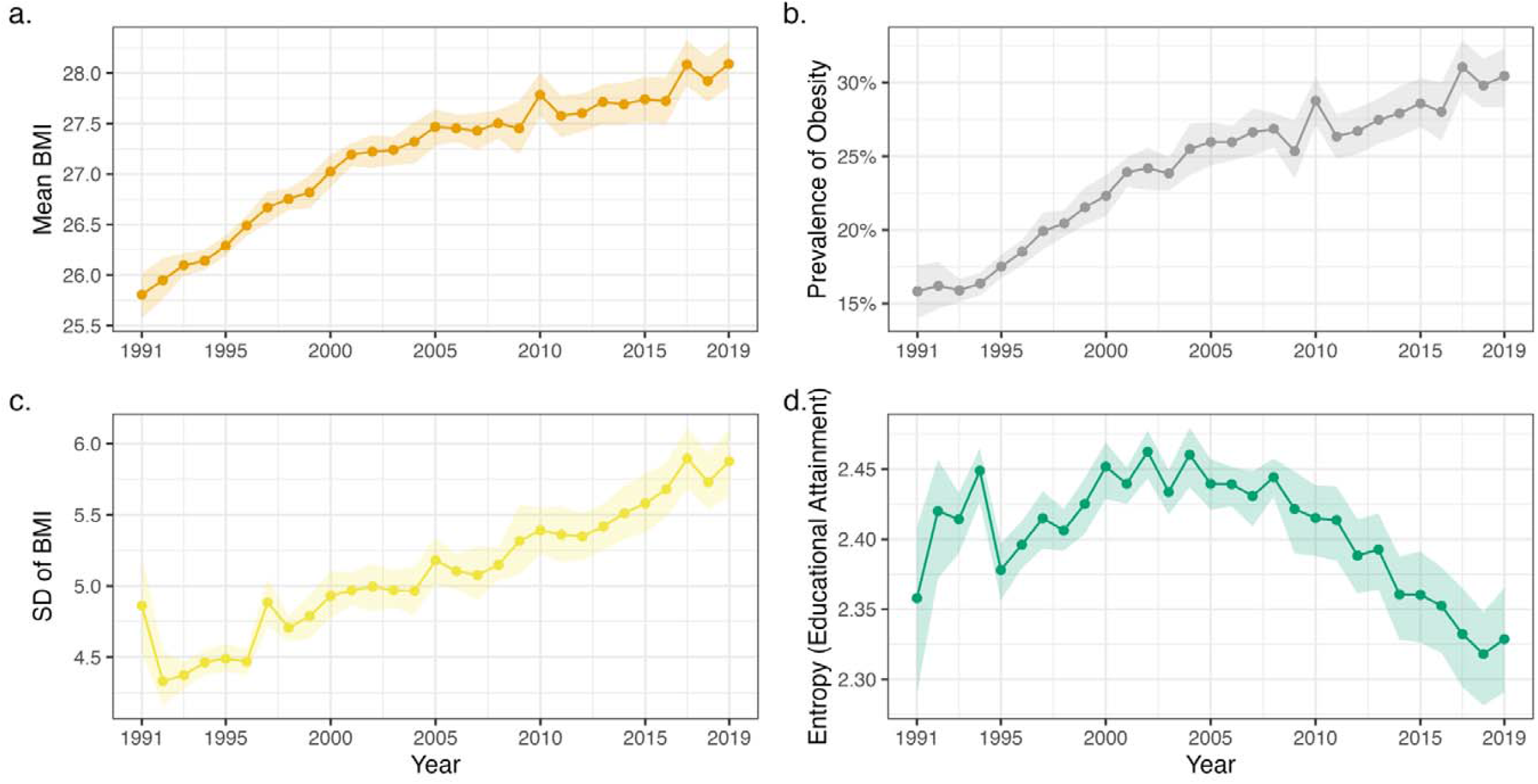
Descriptive statistics (+ 95% confidence intervals) by survey year. (a) Mean body mass index. (b) Proportion of individuals who are obese (BMI ≥ 30 kg/m^2^). (c) Standard deviation of BMI. (d) Shannon’s entropy (a measure of variability) for categorical educational attainment variable. All figures are weighted. Confidence intervals derived using Rao & Wu bootstrap method to account for complex survey design.

### Predicting BMI

Mean BMI increased among all education groups and IMD quintiles across the survey period, including among those with the highest SEP (Figures 2a-b) – for instance, predicted mean BMI increased for the most highly educated group (NVQ 4/5) from 26.2 kg/m^2^ (95% CI = 25.6, 26.7) in 1991 to 28.1 kg/m^2^ (27.7, 28.5) in 2019. More disadvantaged SEP was generally related to higher BMI and there was some evidence that social inequalities widened over time. The difference between the lowest and highest educated groups was 1.0 kg/m^2^ (0.4, 1.6) in 1991 and 1.5 kg/m^2^ (0.9, 1.8) in 2019, while the difference between individuals in the most and least deprived neighbourhoods was 0.5 kg/m^2^ (0.3, 0.8) in 2001 and 1.4 kg/m^2^ (0.8, 1.9) in 2019 (see Supplementary Figure S3 for smoothed results). The trend cannot be explained by changes in age composition over time – generating effect sizes using the age structure of the 2019 HSE sample similar results (results available on request).

**Figure 2:**
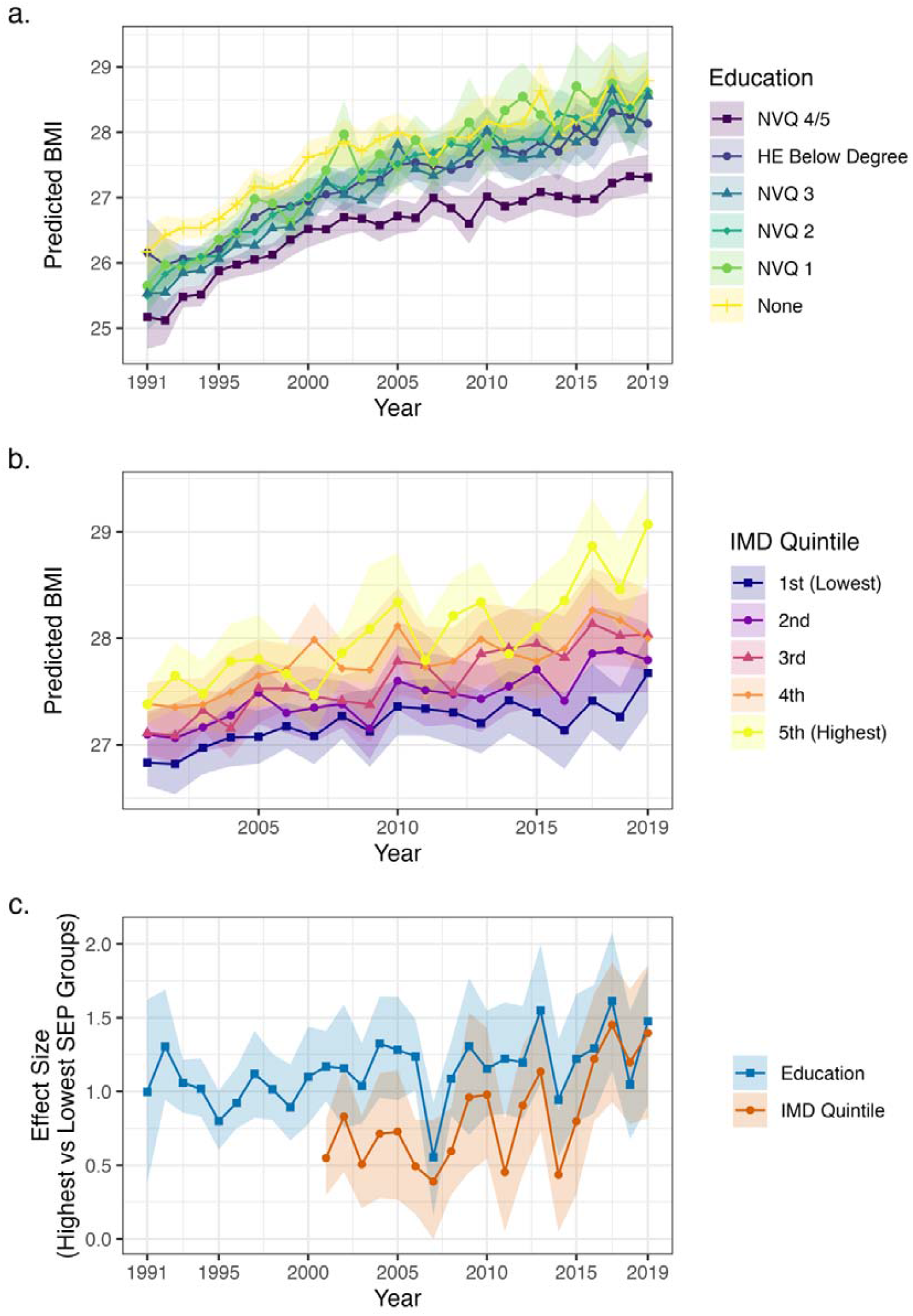
Results of random forest models predicting BMI by survey year. (a) Predicted mean population BMI assuming all individuals have given educational attainment. (b) Predicted mean population BMI assuming all individuals from areas in given IMD quintile. (c) Difference in mean BMI at the population level between highest (NVQ 4/5 or 1^st^ quintile IMD) and lowest (no qualifications or 5^th^ quintile IMD) SEP groups. Confidence intervals calculated using bootstrap samples accounting for complex survey design (500 bootstraps, centile method)

While average BMI increased within SEP groups, so did its variability (Supplementary Figure S4). Given this increasing variability, the total prediction error increased over time, regardless of model used (Figure 3a). In 1991, using age, sex, and education level to predict BMI generated an average prediction error (the difference between predicted and observed BMI) of 3.4 kg/m^2^ (3.2, 3.6). In 2019, prediction error increased to 4.4 kg/m^2^ (4.2, 4.6). Using age, sex, and IMD level, instead, average prediction errors were 3.8 kg/m^2^ (3.7, 3.8) in 2001 and 4.4 kg/m^2^ (4.3, 4.6) in 2019.

**Figure 3:**
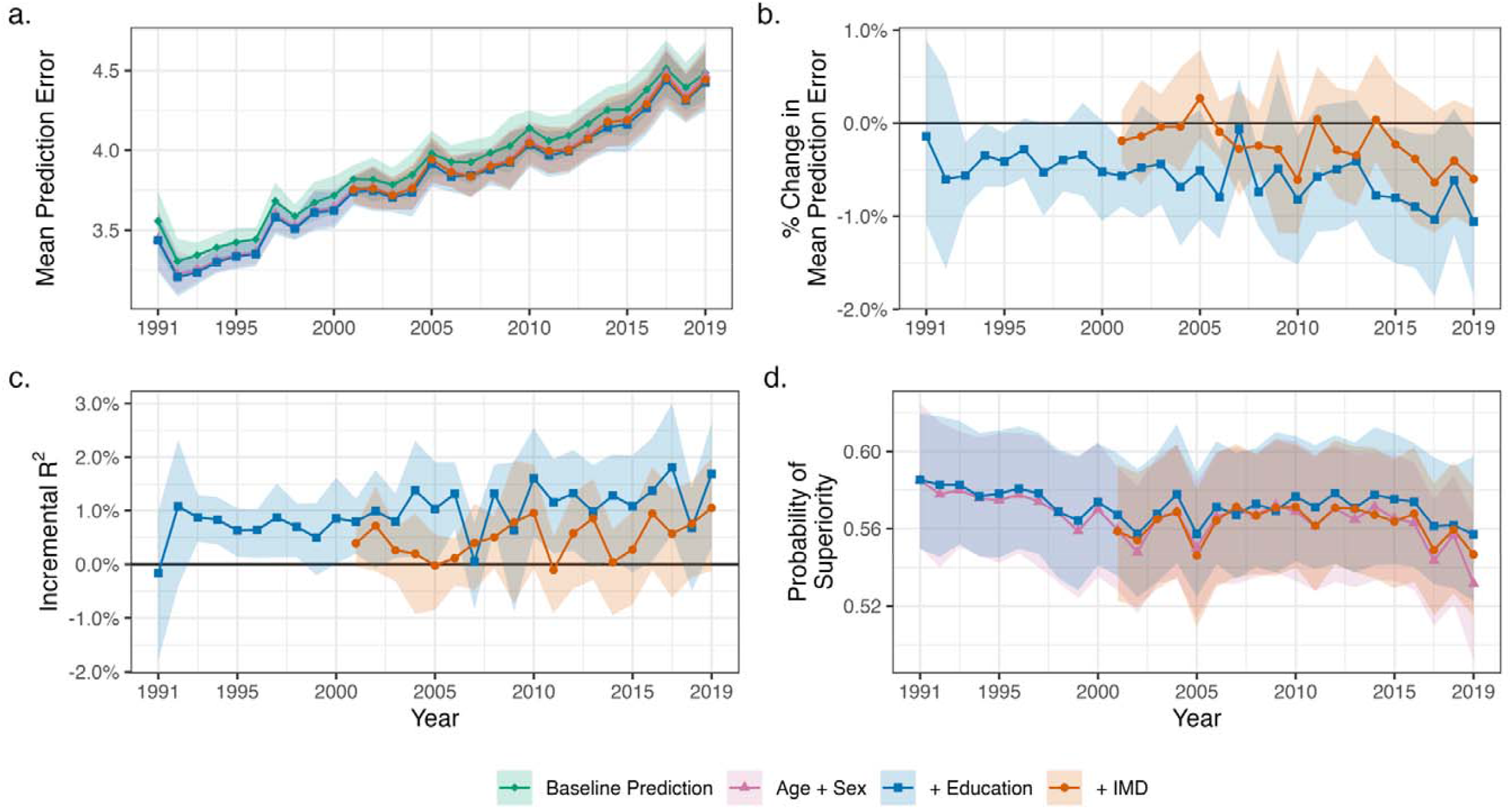
Predictive accuracy of random forest models predicting individuals’ BMI by survey year. (a) Mean absolute error of model predictions by model (i.e., average difference between predicted and observed BMI; baseline prediction uses sample mean, other estimates are random forest models including stated covariates). Higher values are indicative of less accurate prediction. (b) Percentage reduction in prediction error when further including educational attainment or IMD in random forest model (compared to model including age and sex). (c) Incremental R2 when further including educational attainment or IMD in random forest model (compared to model including age and sex). (d) Probability of superiority by model.

While prediction errors increased in absolute size, there was some evidence that education and IMD explained a greater proportion of variation in BMI from 1993 onwards, as measured as the proportion of prediction error reduced by including education or IMD in the random forest model or, alternatively, by incremental R^2^ (Figure 3b-c; see Supplementary Figure S3 for smoothed results). The improvement in prediction attributable to education was 0.14% (−0.9, 1.08) in 1991 and 1.06% (0.17, 1.84) in 2019 (Figure 3b). (A trend of increasing predictive accuracy improvement from including education in models was also observed using data from 1991-2014 only.) Across the studied period, the total reduction in prediction error when including education or IMD in models was very small – less than 1.1% each year (see Supplementary Figure S5 for model residuals). Equivalently, incremental R^2^ was low; for education, 0.83% (0.37, 1.25) in 1994 and 1.69% (0.31, 2.65) in 2019 (Figure 3c). Highlighting this, the ability of education and IMD to distinguish pairs of individuals at higher BMI levels was also generally poor. The probability of superiority derived from models including SEP was 0.59 or lower in each year – little different from the probability of superiority derived from models just including age and sex (Figure 3d).

### Further Analyses

Qualitatively similar results were obtained when linear regression was used instead of the random forest algorithm (results available on request). Qualitatively similar results were also obtained when predicting obesity instead of BMI: social inequalities increased over time as did the proportional improvement in prediction when included SEP in models, but the overall predictive power of SEP was low (see the Supplementary Information Results S1 for full detail). Larger social inequalities were found among women when stratifying the BMI analysis by sex (Supplementary Figure S6). Population level differences in mean BMI according to SEP were approximately twice as large among females compared with males. Accordingly, SEP improved individual level predictions to a greater extent among females, though improvements in predictive accuracy remained low. The relative improvement in predictive accuracy across the study period was more clearly observed among females.

## Discussion

### Summary of Results

The results demonstrate an increase in mean BMI and an increase in the variability of BMI between 1991-2019 in England, as well as an increase in the prevalence of obesity. Mean BMI and prevalence of obesity increased across all education groups and IMD quintiles, and there was an increase in social inequalities over time. However, variability in BMI *within* SEP groups also increased. Whilst the ability of education and IMD to explain the between-person variability of BMI increased over the study period, explained variance remained low and absolute prediction errors increased in size. A similar pattern of results was found when attempting to predict obesity. SEP further had limited utility in identifying, among pairs of individuals, the person with obesity or a higher BMI. Effect sizes were larger in females than males.

### Explanation of Findings

These results are consistent with previous studies showing limited explanatory power of SEP for BMI [12,16–18] and accord with studies showing increased variance within SEP groups over the obesity epidemic [2,18,29–32]. More generally, they are also consistent with findings that shared environmental factors explain limited variance across a wide range of behavioural and health-related traits (the “gloomy prospect” of behaviour genetics [23–27]), as well as with the results of a mass scientific collaboration study showing that socioeconomic outcomes are largely unpredictable even using rich longitudinal survey data [28]. Researchers in one study were able to predict 60% of the variance in BMI among older adults using deep learning methods and detailed socioeconomic, demographic, and other study data (> 450 variables) [46]. However, their analysis also included several variables directly related to health, such as healthcare utilization. Intriguingly, the observed small change in the proportion of variance explained by SEP as group level BMI differences have increased is consistent with a model in which the effects of risk factors for high BMI have uniformly increased in strength over the obesity epidemic [47] – genetic effects have similarly increased, while heritability has remained almost stable [22,48,49].

Our results raise the question of why such low explanatory power of SEP is observed. One reason is that low SEP is neither a necessary nor sufficient cause of high body weight. Instead, SEP is expected to operate distally at the end of long causal chains, the steps of which may be blocked, amplified, or attenuated in the presence or absence of other exposures. For instance, at a population level, neighbourhood deprivation may lead to higher BMI by influencing physical activity via affecting walkability [50], but some individuals may compensate by travelling to surrounding areas or may get sufficient exercise if they do physically demanding jobs. The effects of SEP on BMI may thus be heterogeneous, a process that would entail greater BMI variance within lower SEP groups, which is observed in practice [2,51]. Further, extremely strong effect sizes – stronger than those found in typical epidemiological studies – are required to obtain good predictive power at the individual level [52]. As such, while SEP had an increasingly large effect size on BMI across time, it was not sufficiently large to yield accurate predictions at the individual level.

Our results may have implications for efforts to tackle obesity rates. Assuming the link between SEP and BMI is causal (an assumption supported in some, but not all, quasi-experimental studies; [53– 55]), our results suggest that reducing the social gradient in BMI could reduce but not reverse the obesity epidemic: consistent with other work [2], our results show that obesity rates have increased among all social groups while inequalities within these groups have also increased over time. As has been previously argued, the increasing variability of BMI could mean a one-size-fits-all approach may not be effective as increased variability may reflect distinct determinants [56]. We should, however, note that predicting the effects of intervening on SEP or its mediating pathways is challenging, and it is possible that inequality itself could increase obesity rates [57].

Despite an increasing association between SEP and BMI at the population level, the results suggest limited utility of the use of SEP indicators in predictive algorithms for obesity or BMI. Algorithms to predict obesity based on high-level SEP data are likely to have an unacceptably low sensitivity and specificity – focusing only on those with low SEP would miss the majority of cases. Including SEP in models may be justified for health equity reasons, however [58]; without its inclusion, risk will be systematically underestimated for low SEP individuals.

While SEP does not explain much of the between-person variation in BMI, determining its predictive ability is important as it can motivate the development of more complex and specific theories and highlight the need for other non-standard but highly predictive data. Genetic data are increasingly available – polygenic scores for BMI now achieve R^2^ of 15% [59] – but text or other ‘big’ data could also be useful. A recent study mining the content and style of essays written at age 11 explained almost 20% of the variability in childhood cognitive ability [60], though the ability to predict BMI and other physical health measures is unlikely to be this high.

### Strengths and Limitations

Strengths included objective measurement of BMI and use of data spanning almost three decades of the obesity epidemic in England. We examined measures of individual and area-level SEP, measures that are easy to collect (and thus may appear in predictive algorithms) and have been widely studied in the social inequality literature previously. Nevertheless, these variables were relatively high level and restricted to a small number of categories, limiting potential predictive accuracy. The measures were also based on current SEP; life course measures of SEP – or of body weight (e.g., ever obese) – would have yielded more accurate predictions (though the gloomy prospect makes us circumspect as to the degree of improvement). Improvements in predictive accuracy may also have been greater if covariates other than age and sex were included in models, as this would allow for the determination of more granular interaction effects. The data we used were also cross-sectional. Assuming that our estimates at least partly confounded [see, e.g., 61], we are likely to have obtained optimistic estimates of predictive accuracy, relative to intervening directly on SEP [62]. Finally, the random forest models may have been too flexible and overfit the data, producing poor out-of-sample predictions. Nevertheless, using OLS regression yielded similar results.

### Conclusions

While absolute inequalities in BMI and obesity according to education and neighbourhood deprivation increased in England between 1991 and 2019, within group inequalities also increased and were large relative to between groups inequalities, contributing to the weak explanatory power of SEP. Though explanatory power increased over the study period, it remained low which suggests that reducing inequality is unlikely to reverse the large impact on the obesity rates which increased across all SEP groups since the beginning of the obesity epidemic. Nevertheless, the possibility of heterogeneous effects of SEP means that targeted attention within SEP groups could be fruitful.

## Data Availability

Health Survey for England data are available via the UK Data Service (https://ukdataservice.ac.uk/). The code used to run the analysis is available at https://osf.io/smd7z/.

https://osf.io/smd7z/

## Statements

### Declaration of interest

All authors declare no conflicts of interest.

### Funding

The funders had no final role in the study design; in the collection, analysis, and interpretation of data; in the writing of the report; or in the decision to submit the paper for publication. All researchers listed as authors are independent from the funders and all final decisions about the research were taken by the investigators and were unrestricted. DB and RS are supported by the Economic and Social Research Council (grant number ES/M001660/1); DB and LW by the Medical Research Council (MR/V002147/1). CBS is funded by the ERSC [ES/V012789/1] and the National Institute of Health Research [COV-LT-0009-28654].

### Acknowledgements Author contributions

All authors contributed to and agreed upon the analysis plan. LW carried out the analysis and wrote the first draft of the manuscript. All authors provided critical revisions.

## Notes

### Competing Interest Statement

The authors have declared no competing interest.

### Author Declarations

Health Survey for England data are available via the UK Data Service (https://ukdataservice.ac.uk/).

