## Supplemental Information for "Trends across time in socioeconomic differences in body mass index: a comparison of population and individual-level approaches"

Supplementary Information

### Results S1

#### Predicting Obesity

As obesity is of particular research and policy interest, we repeated the random forest model analyses using obesity as the outcome measure. For this analysis, the measures of predictive accuracy were the Brier score and the area under the receiver operating curve (AUC). For binary variables, these are analogous to root mean square error and probability of superiority, respectively.

The prevalence of obesity increased among all education groups and IMD quintiles (Figure S7a-b). The difference between the prevalence of obesity for those with no qualifications and those with degree level qualifications or above increased from 7.49% (95% CI = 3.36%, 11.5%) in 1991 to 11.3% (6.75%, 14.3%) in 2019 (Figure S7c; see also Supplementary Figures S8). The corresponding difference between individuals in the most and least deprived neighbourhoods was 4.8% (2.84%, 6.76%) in 2001 and 9.84% (5.88%, 13.1%) in 2019 (Figure S7c; see also Supplementary Figures S8). Again, the results cannot be explained by changes in age composition over the study period (results available on request).

As with BMI, prediction error – as measured by the Brier score - increased over time (Figure S9a). While the explanatory power of education and IMD increased overall over the study period, the total reduction in prediction error when including the variables was small – less than 1.5% points in each year (Figure 9b; see also Supplementary Figures S8). Supplementary Figure S10 shows the difference in the average predicted probability of obesity between those who were obese and those who were not – as can be seen, there is little separation according to observed obesity. Consequently, the ability of education or IMD to distinguish pairs of individuals was low: the AUC statistics were less than 0.605 in each year (Figure S8d), little difference from the AUC in models including just age and sex.

### Figures

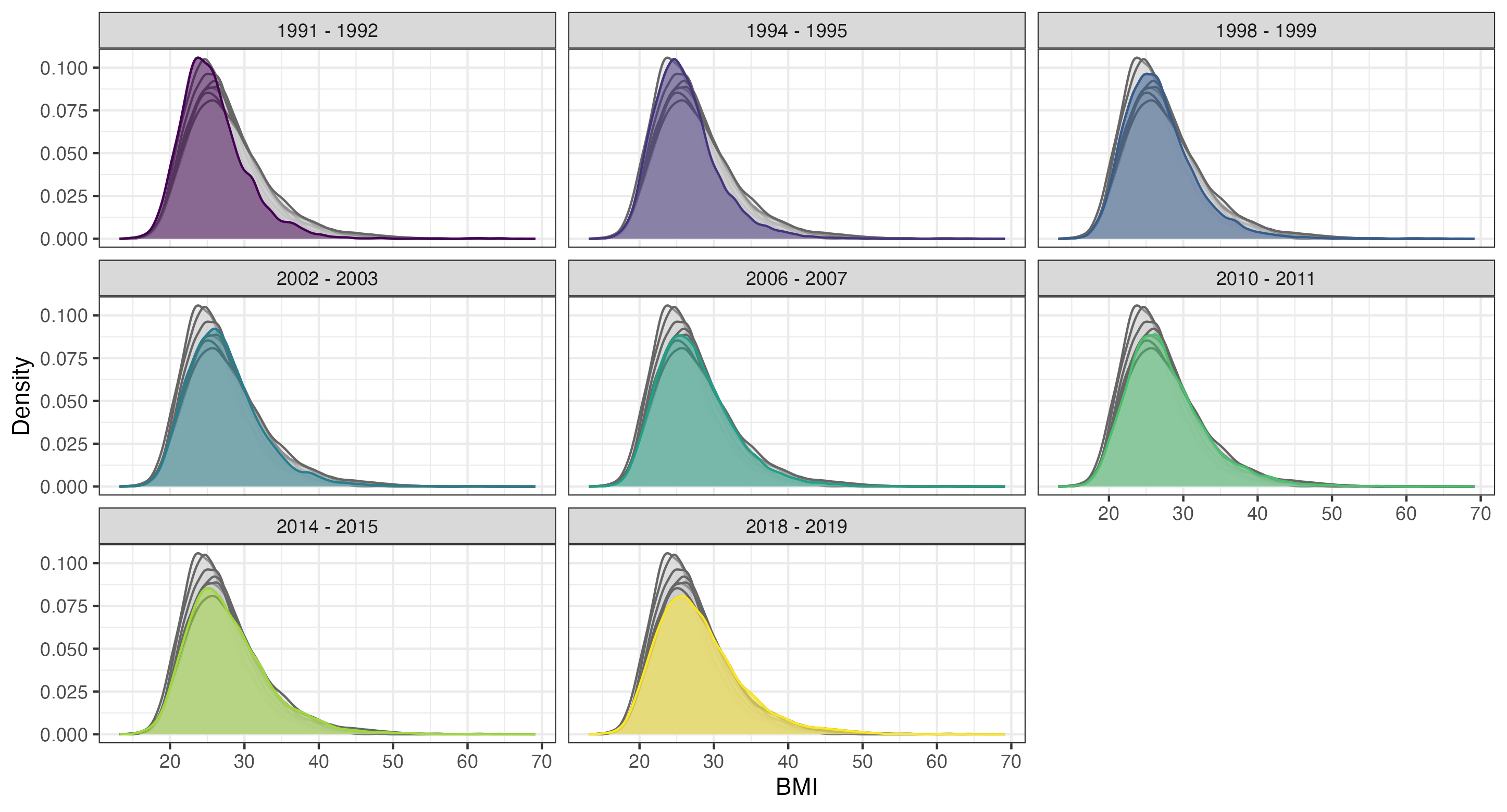

Figure S1: Distribution of BMI by year (two years grouped together), Health Survey for England. Survey weighted figures. In each plot, coloured areas show the distribution of BMI in the stated survey years. Grey areas show the distribution of BMI in the other survey years and are provided for comparison.

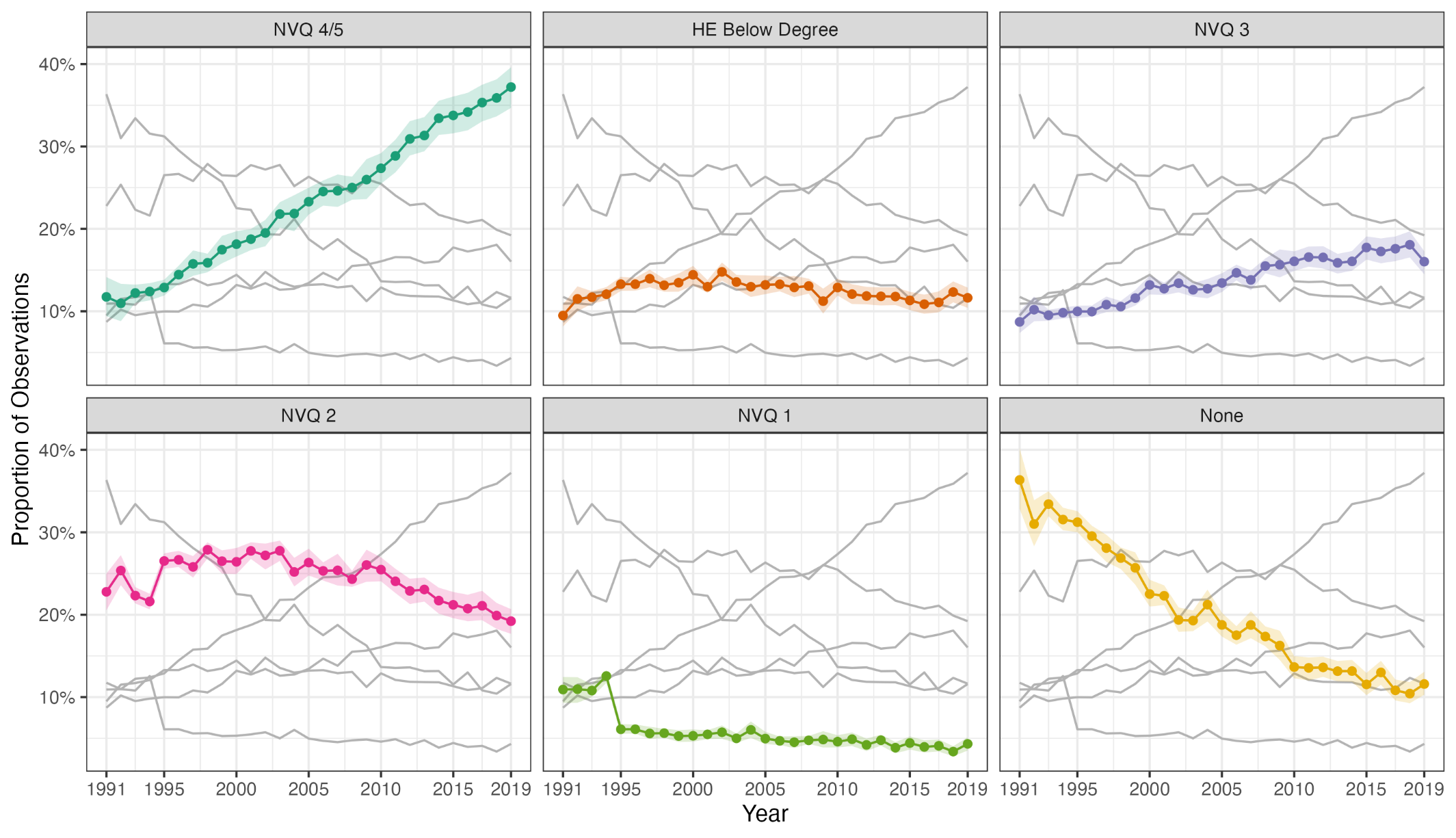

Figure S2: Proportion in each educational category by year, Health Survey for England. Survey weighted figures. Confidence intervals derived using Rao & Wu bootstrap method to account for complex survey design.

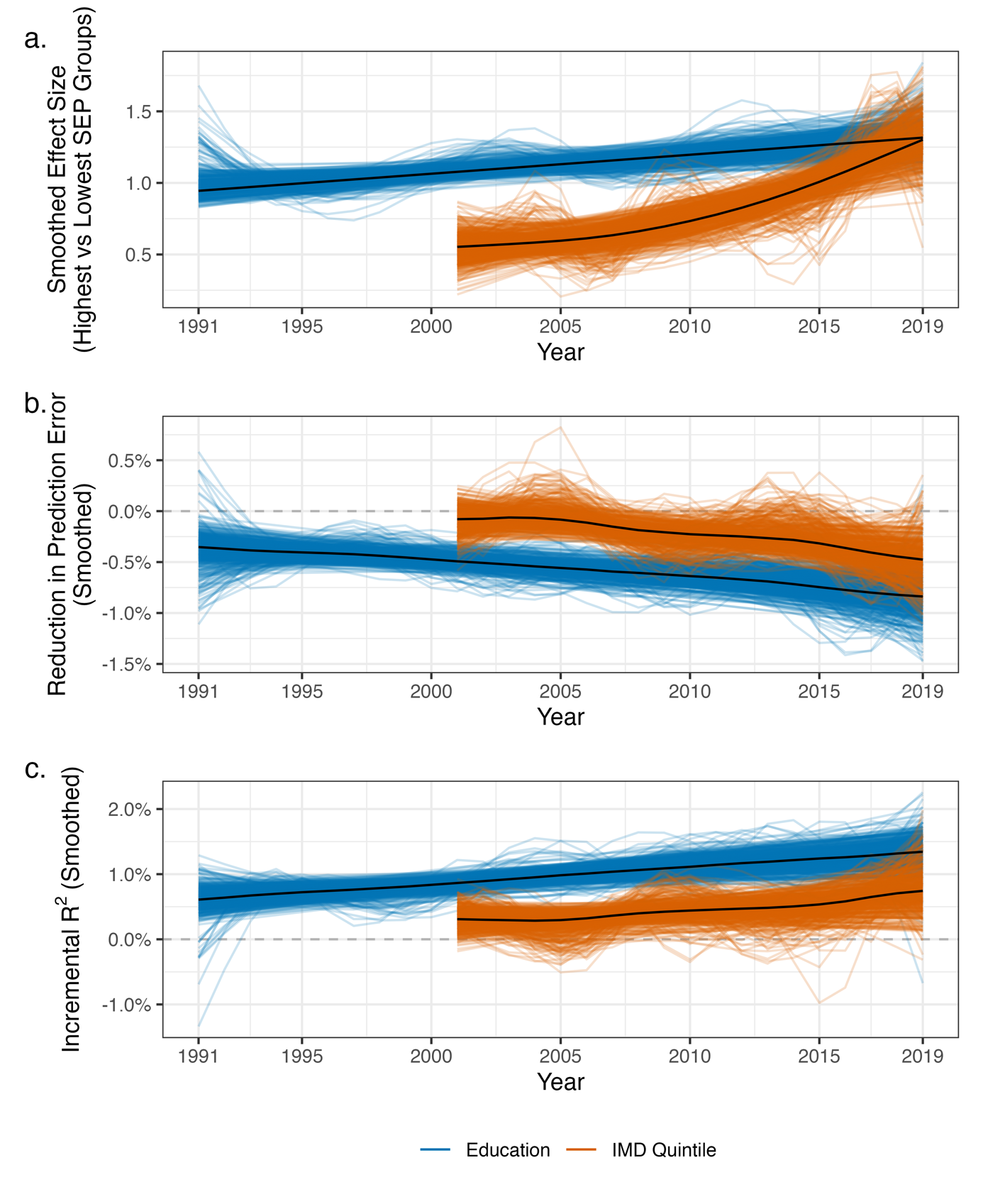

Figure S3: Smoothed results of random forest models predicting BMI (each line is the result from a single bootstrap sample). (a) Difference in mean BMI between highest and lowest SEP groups (NVQ 4/5 vs. no qualifications; or least vs most deprived IMD quintile). (b) Percentage reduction in prediction error when further including educational attainment or IMD in random forest model (compared to model including age and sex). (c) Incremental R^2^ when further including educational attainment or IMD in random forest model (compared to model including age and sex). Smoothing achieved by regressing estimate on year using generalized additive model, performed for each bootstrap sample separately.

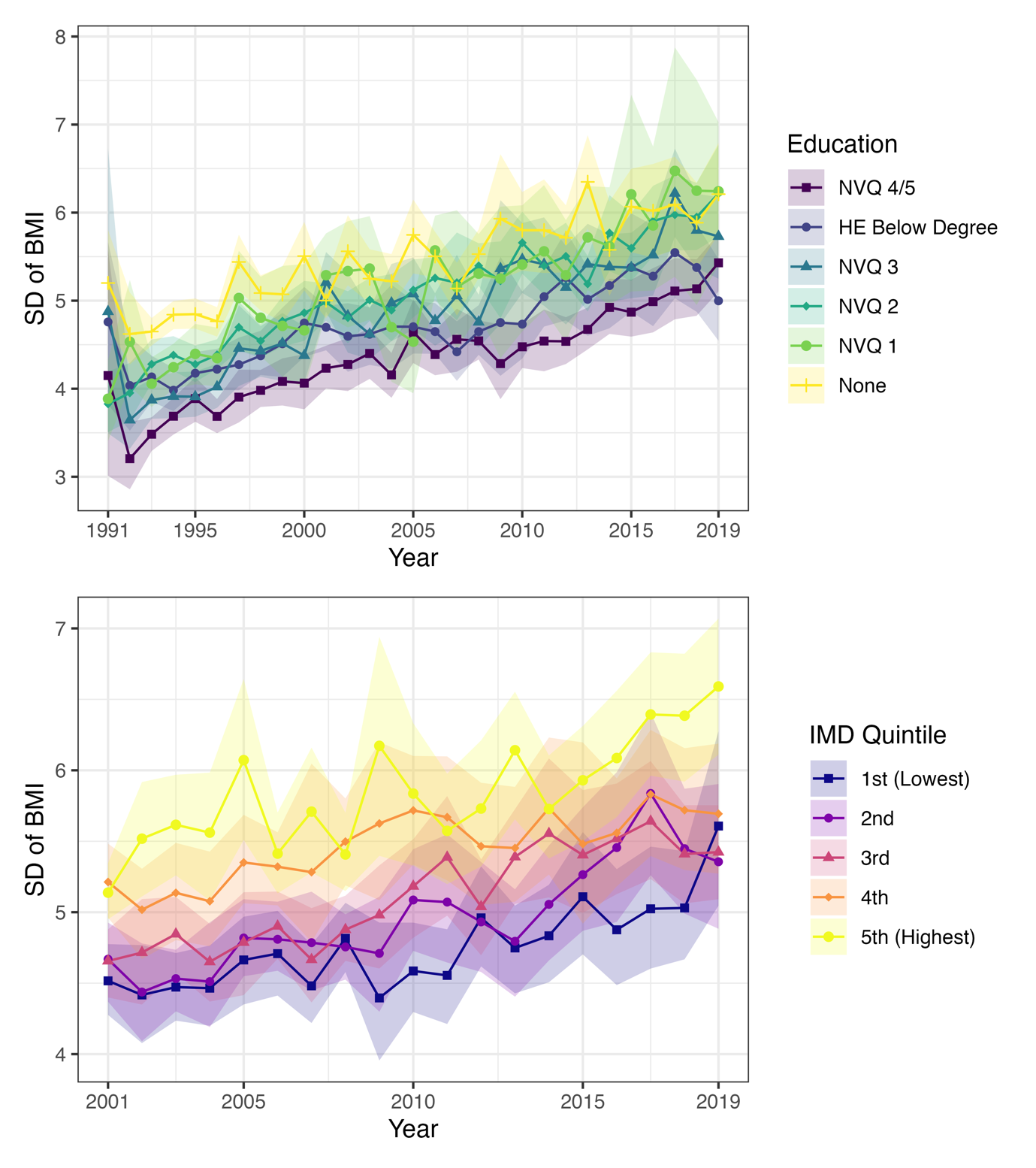

Figure S4: Standard deviation (SD) of BMI within each SEP group by survey year. To account for changes to age and sex distribution within each SEP groups, figures calculated by running random forest model of BMI on age and sex, extracting model predictions and calculating residuals (i.e., observed BMI – predicted BMI), and then calculated SD of the residuals within each SEP group. Confidence intervals derived using Rao & Wu bootstrap method (centile method) to account for complex survey design.

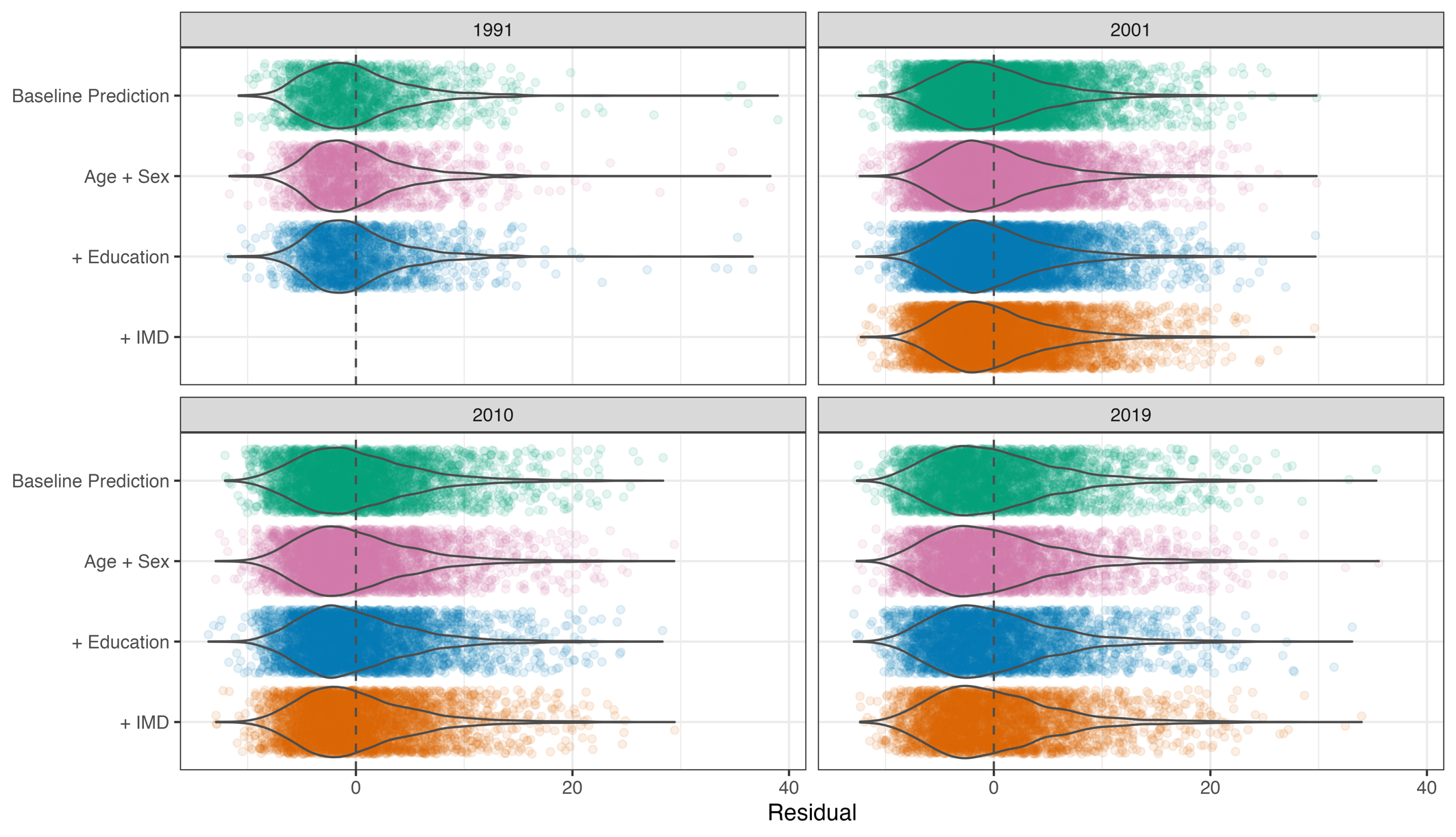

Figure S5: Distribution of prediction errors by year and model. Years 1991, 2000, 2010, 2019 chosen to simplify presentation. Note, these residuals are from the same sample that was used to generate the predictions. They are thus likely to be biased by overfitting.

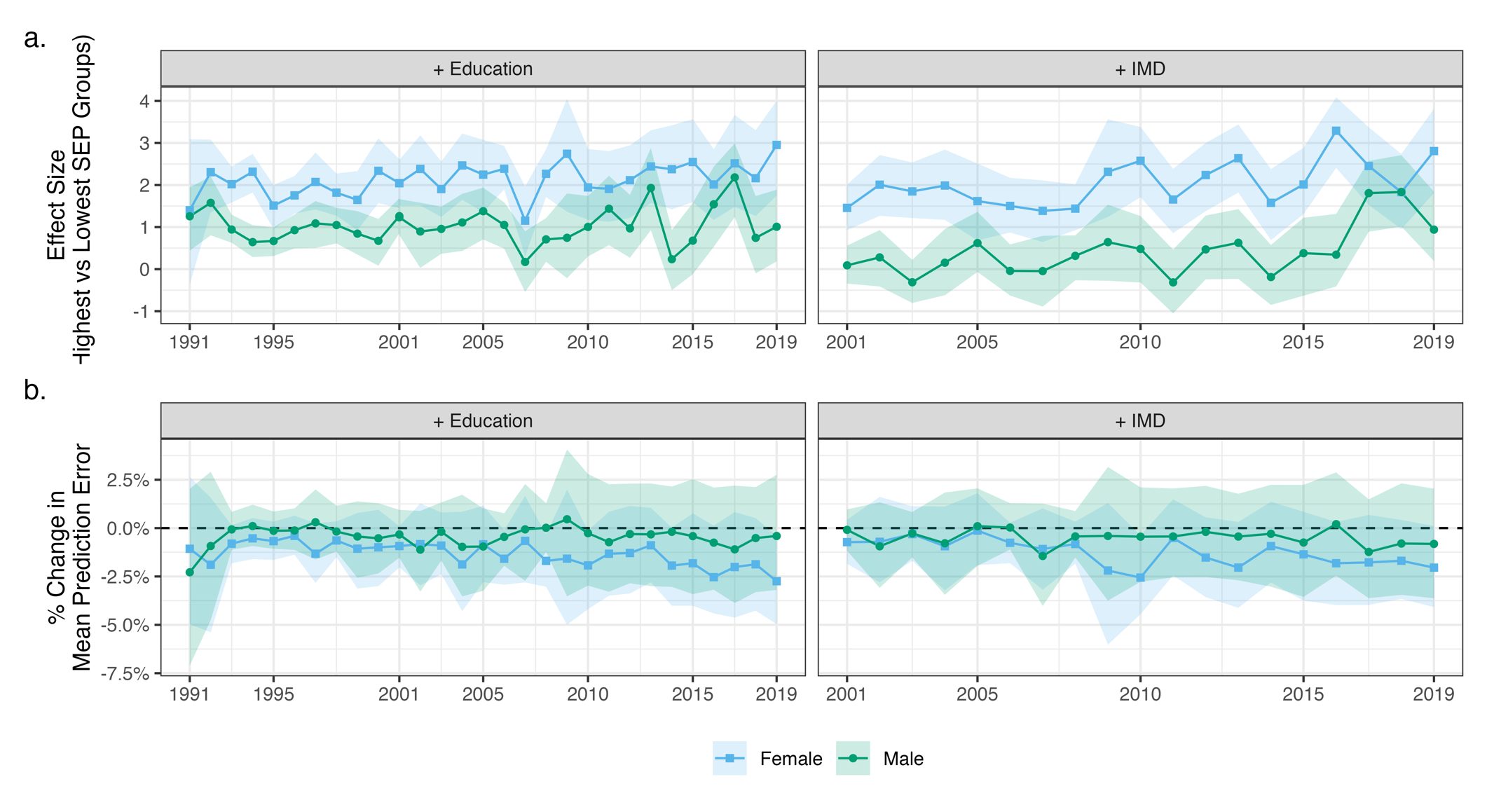

Figure S6: Results of random forest models predicting BMI by survey year and sex. (a) Difference in mean BMI at the population level between highest (NVQ 4/5 or 1st quintile IMD) and lowest (no qualifications or 5th quintile IMD) SEP groups. (b) Percentage reduction in prediction error when further including educational attainment or IMD in random forest model (compared to model including age). Confidence intervals calculated using bootstrap samples accounting for complex survey design (500 bootstraps, centile method)

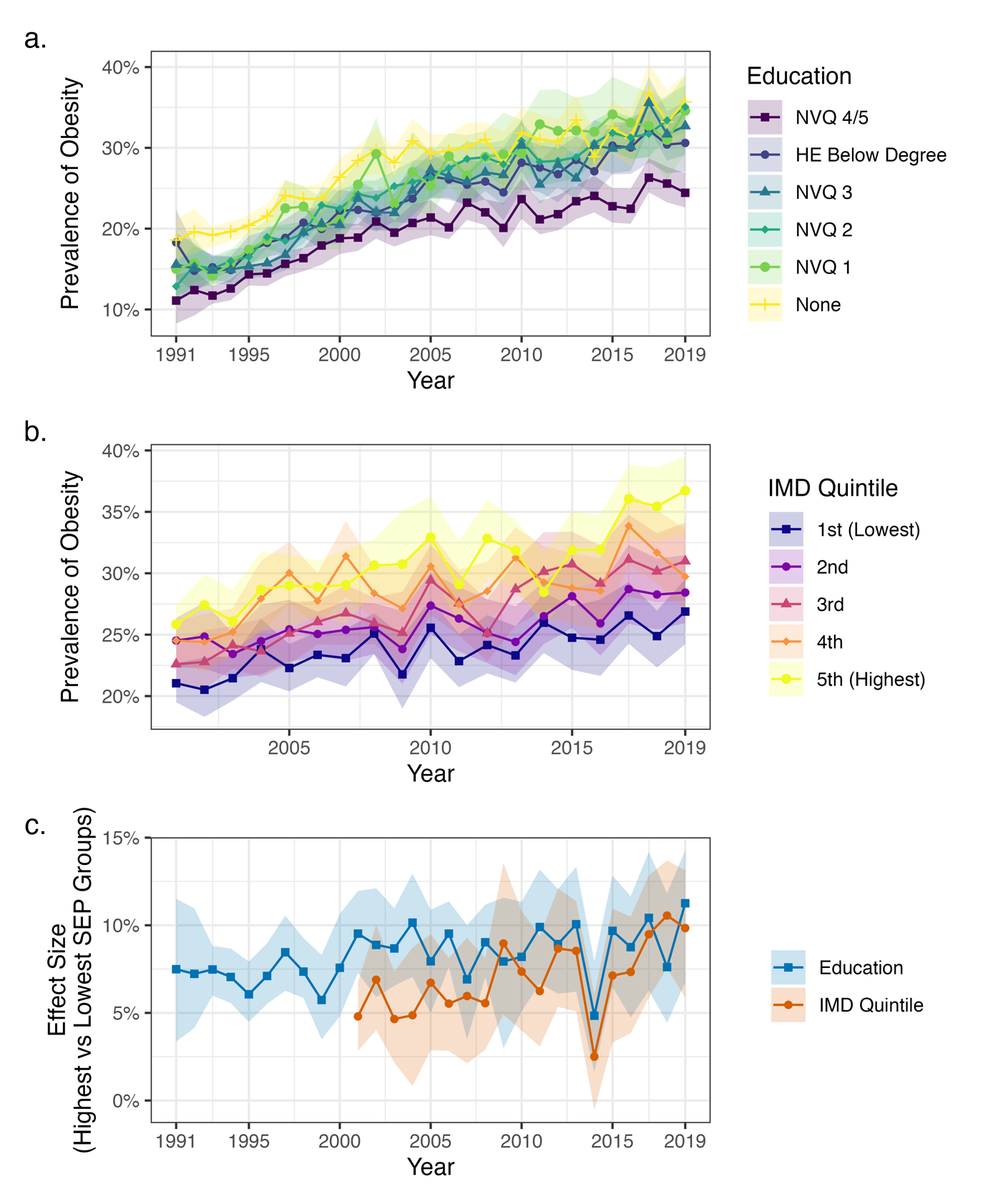

Figure S7: Results of random forest models predicting (probability of) obesity by survey year. (a) Predicted prevalence of obesity assuming all individuals have given educational attainment. (b) Predicted prevalence of obesity assuming all individuals from areas in given IMD quintile. (c) Difference in prevalence of obesity between highest (NVQ 4/5 or 1st quintile IMD) and lowest (no qualifications or 5th quintile IMD) SEP groups. Estimated from random effects. Confidence intervals calculated using bootstrap samples accounting for complex survey design (500 bootstraps, centile method).

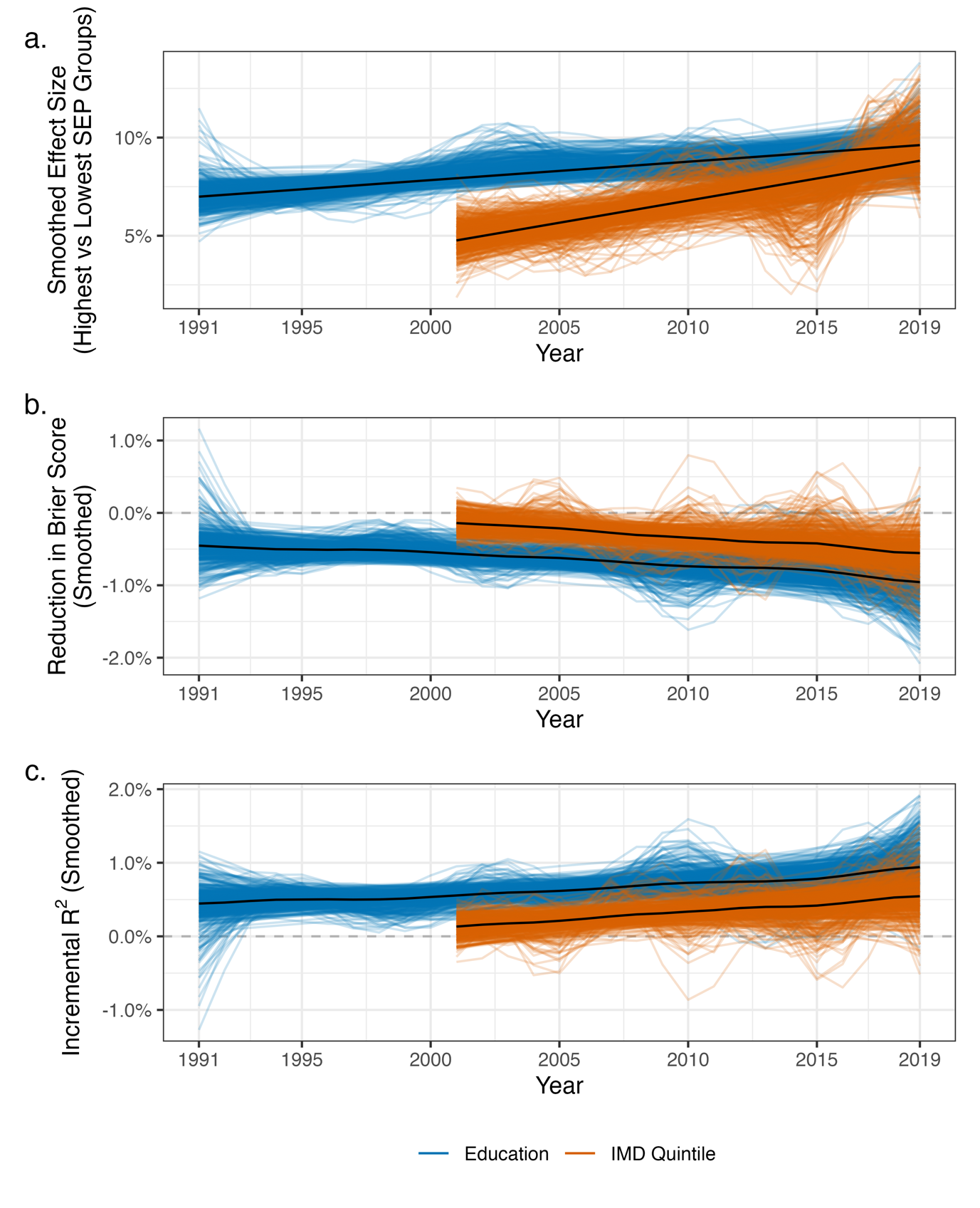

Figure S8: Smoothed results of random forest models predicting obesity. (a) Difference in mean BMI between highest and lowest SEP groups (NVQ 4/5 vs. no qualifications; or least vs most deprived IMD quintile). (b) Percentage reduction in Brier score when further including educational attainment or IMD in random forest model (compared to model including age and sex). (c) Incremental R^2^ when further including educational attainment or IMD in random forest model (compared to model including age and sex). Smoothing achieved by regressing estimate on year using generalized additive model, performed for each bootstrap sample separately.

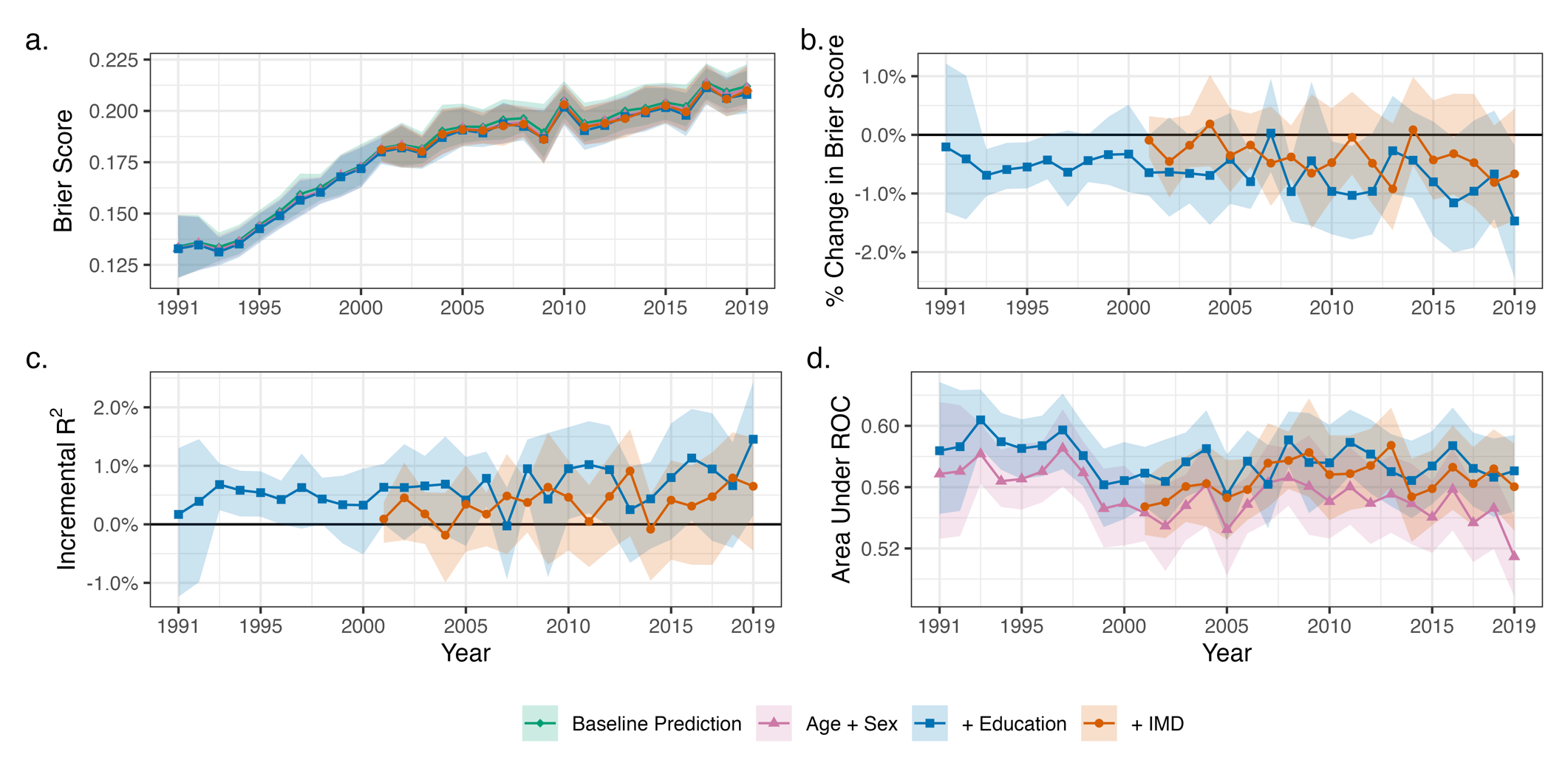

Figure S9: Results of random forest models predicting (probability of) obesity by survey year. (a) Root mean square error (Brier score) of model predictions by model (baseline prediction uses sample mean; other estimates are random forest models including stated covariates). (b) Percentage reduction in prediction error when further including educational attainment or IMD in random forest model (compared to model including age and sex). (c) Incremental R^2^ when further including educational attainment or IMD in random forest model (compared to model including age and sex). (d) Area under the ROC - probability a randomly chosen obese individual has higher predicted probability of obesity than randomly chosen non-obese person by model.

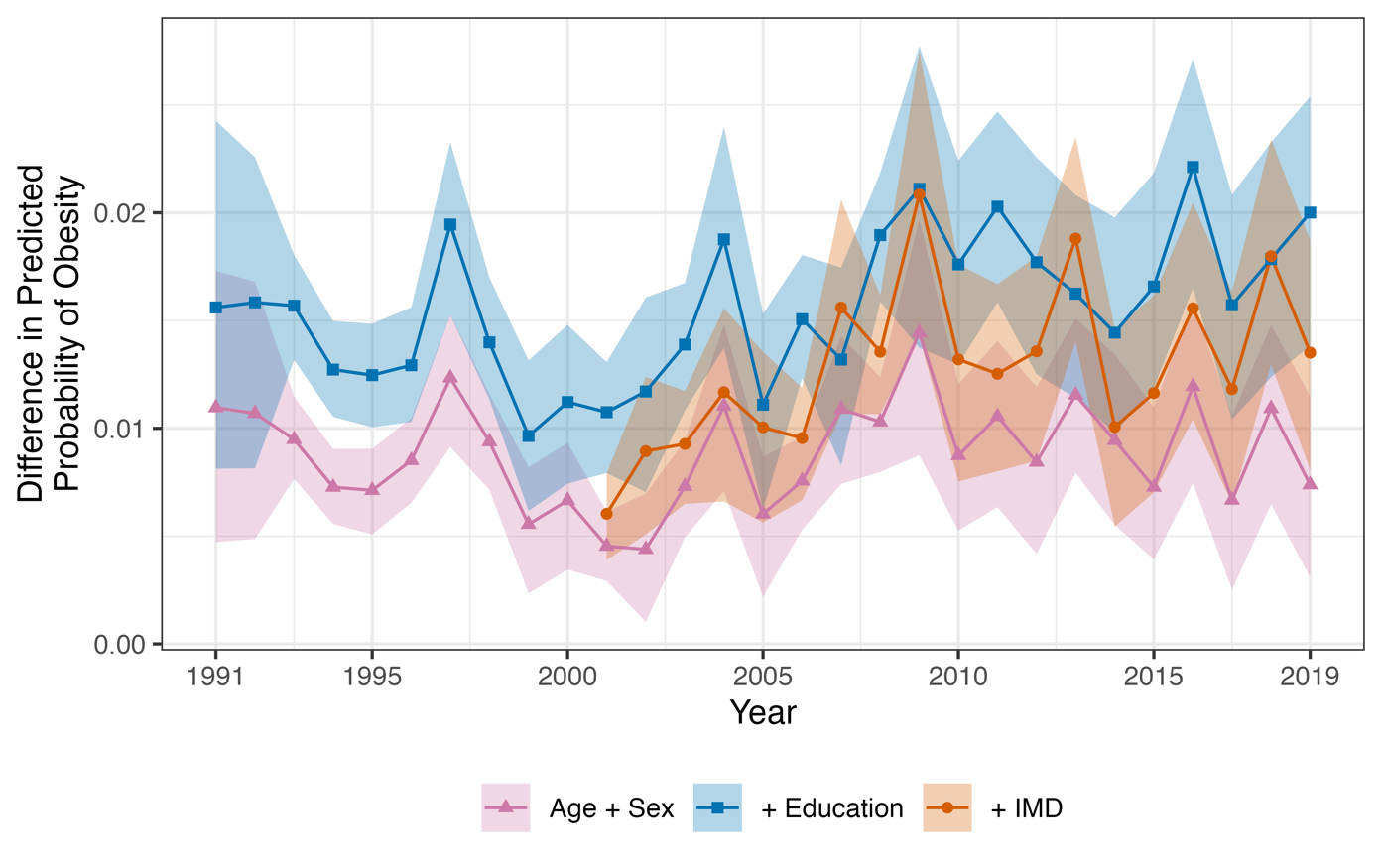

Figure S10: Difference in the average predicted probability of obesity between obese and non-obese participants by survey year and model. Derived from random forest models. Color indicates covariates included in the random forest model. Confidence intervals calculated using bootstrap samples accounting for complex survey design (500 bootstraps, centile method).
